# Bacteriological Profile and Antimicrobial Susceptibility Patterns of Uropathogens at a Tertiary Care Hospital in North India: A Retrospective Descriptive Study

**DOI:** 10.64898/2026.08.02.26359083

**Authors:** Girlie Tambirai Mapere, Anshu Kumar Singh, Umesh Kumar, Pankaj Kishor Mishra

**Author notes:** **Corresponding author: Anshu Kumar Singh, Address for Correspondence**: Department of Medical Laboratory Science, Subharti College of Allied and Health Sciences, Swami Vivekanand Subharti University, Meerut, Uttar Pradesh, India.

## Abstract

**Background:** The main cause of urinary tract infections (UTIs) are Gram negative bacteria with *Escherichia coli* as the leading cause and other important pathogens such as *Klebsiella pneumoniae, Pseudomonas aeuriginosa* and *Enterococcus faecalis*. Over the years uropathogens have become resistant to commonly used antibiotics, including penicillin’s, cephalosporins and fluoroquinolones. Antimicrobial resistance (AMR) in UTIs is mainly caused by the misuse and overuse of antibiotics, recurrent infections, and healthcare-associated factors such as catheterization.

**Objectives:** The aim of this study was to describe the bacteriological profile and antimicrobial susceptibility patterns of uropathogens isolated from positive urine cultures at Chhatrapati Shivaji Subharti Hospital, Meerut, a tertiary care centre in North India and develop an institutional antibiogram to support empirical prescribing and antibiotic stewardship at this institution.

**Materials & Methods:** The study analysed 50 positive urine culture samples and their antimicrobial susceptibility records from July 2025 to December 2025. The isolates were identified, and antimicrobial susceptibility testing was performed using the disc diffusion method and automated Biomerieux Vitek 2 Compact machine. The collected data was analysed using descriptive statistics and Fisher’s exact test.

**Results:** Gram-negative bacteria accounted for 80.0% (40/50) of the culture-positive urine isolates. *Escherichia coli* was the most frequently isolated uropathogen (n=23, 46.0%), followed by *Klebsiella pneumoniae* (n=12, 24.0%), *Candida* spp. (n=6, 12.0%), *Enterococcus* spp. (n=4, 8.0%), *Pseudomonas aeruginosa* (n=3, 6.0%), and *Enterobacter cloacae* (n=2, 4.0%). Of the total, 74% (37/50) of isolates came from Inpatient samples. *E. coli* had a 100% resistance to ampicillin and ceftriaxone, 95.7% to ciprofloxacin and cefepime, and 73.9% to meropenem, with fosfomycin (86.4% sensitive) and colistin (69.6% sensitive) as the only effective antimicrobials. *K. pneumoniae* had 100% resistance to ceftriaxone, amoxicillin-clavulanate, and piperacillin-tazobactam; carbapenem resistance ranged from 83.3% to 91.7%, and colistin was the only consistently effective treatment (83.3% sensitive). Of all the 35 tested *Enterobacteriaceae* isolates, Extended-Spectrum Beta-Lactamases (ESBLs) positivity was 100% with Carbapenem-Resistant Enterobacterales (CRE) positivity at 85.0%. All the bacterial isolates met Multi-drug Resistant (MDR) criteria. 31 of 35 (88.6%) tested *Enterobacteriaceae* isolates showed ESBL and CRE co-positivity. The six strains of *Candida* demonstrated total sensitivity to all the antifungal drugs used.

**Conclusion:** There is a critical burden of AMR at this hospital with 100% ESBL positivity, 85% CRE, and 100% MDR among all bacterial isolates. This study provides the first baseline institutional antibiogram to guide empirical prescribing and antibiotic stewardship at Chhatrapati Subharti Hospital.

## INTRODUCTION

Urinary tract infections (UTIs) refer to the infection of the urinary tract region of the urethra, bladder, ureters and kidneys by bacteria [1]. While UTIs are known to be manageable, 404.61 million worldwide in 2019 had a UTI and 236,790 deaths were recorded [2]. The risk factors for developing UTIs include age, gender, the use of urinary catheters, longer hospital stays and previous exposure to antimicrobials [3]. Women are mostly affected by UTIs because they have a shorter urethra compared to men which is closer to the rectum that carries waste and bacteria and thus provides a shorter pathway to the bladder [2,4]. Additionally, women experience hormonal changes during their menstrual cycles, pregnancy and menopause making them more at risk [4]. An interesting fact is that when men get UTIs, they tend to be worse than women’s and are often associated with other health problems like prostate enlargement, diabetes, catheter use, or another urinary blockage all of which make it harder to recover [4].

In most healthcare settings, 9.4% of patients are diagnosed with UTIs and are one of the most common healthcare associated infections (HCAIs) with approximately 1-19% prevalence, accounting 30-40% of all HCAIs reported by hospital settings [5]. UTIs occur in both IPD and OPD settings, but their prevalence, types of bacteria causing them and how resistant those bacteria are to antibiotics differs between the two settings. In the OPD, UTIs are mostly community-acquired and caused by fewer antibiotic-sensitive pathogens, making them easier to treat. On the other hand, IPD isolates, include more diverse multidrug-resistant organisms, invasive devices such as urinary catheters, and greater clinical severity [6,7].

As mentioned by Akhavizadegan et al (2021), the types of antibiotics that are not effective on different uropathogens are increasing and this leads to increased failure in treating UTIs. Recently, it has been reported that approximately 700,000 people worldwide die annually due to AMR infections and it has been predicted that this number would reach 10 million by 2050 [8]. There has been a worrying rise in the resistance of uropathogens like *E. coli* and *K. pneumoniae* in India, mainly in South India. Imipenem resistance, for instance, rose from 3.1% to 32.9% between 2011 and 2017 with many antibiotics now exhibiting resistance rates higher than 20% [9].

India has a high disease burden and stands at a critical juncture in the fight against AMR worldwide [10]. The results of this growing crisis keep growing and diminish decades of medical progress, healthcare delivery and potentially hindering the achievement of several Sustainable Development Goals [10].

The bacterial spectrum causing UTIs is different across geographical location, patient age, and whether the infection is community acquired or hospital-acquired [11]. UTIs are caused by gram-negative, gram-positive bacteria as well as some fungi. Although the local epidemiology of the pathogens can be different, it has been noted that *E. coli* causes most of the UTI infections [1]. Other pathogens causing UTIs include *Klebsiella* species, *Proteus* species and *Pseudomonas aeruginosa.* Gram-positive bacteria such as Staphylococcus *aureus*, coagulase-negative *Staphylococcus*, and *Enterococcus* species are also important uropathogens. While bacteria are the main cause of UTIs *, Candida* spp has also been identified as a diagnostic challenge [12].

Although there is research on UTIs and AMR, some gaps restrict the practical use of existing data. Comparisons between IPD and OPD within the same tertiary hospital are still lacking because many studies combine both IPD and OPD cultures or focus on only one group. This makes it difficult to develop setting-specific treatment protocols. Secondly, majority of the studies only focus on antibiotic susceptibility and do not report phenotypic classification such as ESBL, CRE, and MDR. These are clinically important resistance mechanisms that are underreported and without thorough characterization, recommendations on last-resort antibiotics remain incomplete and may pose safety risks. Lastly, while fungal uropathogens especially *Candida* are increasingly becoming clinically relevant, they are often excluded from uropathogen studies which is an important gap since the WHO published its first Fungal Pathogen Priority List in 2022.

These limitations prove the need for studies that go beyond simple prevalence reporting in order to give comprehensive, setting-specific, and mycologically inclusive data, such is the method used in this work. This study addresses this gap by providing the first comprehensive bacteriological audit of culture-positive uropathogens and their antimicrobial susceptibility patterns at Chhatrapati Shivaji Subharti Hospital, Meerut a 938-bed tertiary care centre serving a large North Indian population. This will develop the hospital antibiogram data required to guide evidence-based empirical prescribing, infection control policy, and antibiotic stewardship programme development at this institution.

## MATERIALS AND METHODS

### Study design

This was a retrospective descriptive study carried out at Chhatrapati Shivaji Subharti Hospital in Uttar Pradesh over a six months period from July 2025 to December 2025.

### Study population

The study used patient data from IPD and OPD departments whose urine samples were submitted to the Central Microbiology Laboratory of Chhatrapati Shivaji Subharti Hospital for culture and antimicrobial susceptibility testing. The analysis used a total of 50 culture-positive urine isolates.

### Inclusion criteria

This study included patients aged 18 years and older whose urine was tested positive for pathogens with a bacterial or fungal growth at a colony count of ≥ 10⁵ CFU/ml.

### Exclusion criteria

This study did not take data from urine samples that were contaminated or recorded to have commensal bacteria, showed no growth and duplicate samples from same patient.

### Data collection

The data was collected retrospectively from laboratory registers and the Laboratory Information System (LIS) software of the Central Microbiology Laboratory, Chhatrapati Shivaji Subharti Hospital. The following parameters were taken: patient identifier, age, gender, ward and clinical setting (IPD/OPD), date of sample collection, organism isolated, colony count, Gram stain result, identification method, antibiotic susceptibility profile, ESBL screening result, carbapenemase screening result, and MDR classification. The data was entered into a structured database in Microsoft Excel for analysis.

### Laboratory methods and antimicrobial susceptibility testing

The collection of the urine sample was done under sterile conditions. A colony count of ≥ 10^5^ CFU/ml was considered as significant bacterial count. The samples were first cultured on Cysteine Lactose Electrolyte Deficient (CLED) agar and incubated at 37°C for 24 hours. The colonies that showed growth were characterised by morphology such as mucoid vs non-mucoid, lactose-fermenting vs non-lactose-fermenting, performed Gram staining and microscopy. Biochemical identification was performed using oxidase test. The species and antibiotic susceptibility were identified by Biomerieux Vitek 2 Compact machine. The manual method of antibiotic susceptibility was carried out by the Kirby-Bauer disc diffusion method. The antimicrobial susceptibility test was determined by measuring the zone of inhibition using Clinical and Laboratory Standards Institute (CLSI) interpretive chart for susceptibility tests and the European Committee on Antimicrobial Susceptibility Testing (EUCAST).

The diameters of the zones of complete inhibition were measured using mm of calipers. ESBL screening was performed on *Enterobacteriaceae* isolates only *E. coli and K. pneumoniae* in accordance with standard laboratory protocol, as ESBL production is not a clinically applicable resistance mechanism in *Pseudomonas aeruginosa or Enterococcus spp*.

Antibiotics used for Gram-negative bacteria were Fosfomycin, Amoxicillin-Clav, Ampicillin, Ceftriaxone, Ciprofloxacin, Cotrimoxazole, Nitrofurantoin, Pip-Tazobactam, Amikacin, Cefepime, Colistin, Ertapenem, Gentamicin, Imipenem and meropenem. Antibiotics used for Gram-positive bacteria were Fosfomycin, Ampicillin, Ciprofloxacin, Nitrofurantoin, Tetracycline and Vancomycin, Linezolid, Teicoplanin, Penicillin G and High-Level Gentamicin. Multidrug resistance was defined as the ability to withstand one agent in three or more antimicrobial categories, as per the standardised international definition. ESBL production was confirmed by the combined disc method and automated ESBL detection on the Biomerieux VITEK-2 system while, carbapenemase production was confirmed by the Biomerieux VITEK-2 system.

### Quality Control

Quality control (QC) was done in accordance with CLSI guidelines. The standard QC panel used were *E. coli* ATCC25922, *K. pneumoniae* ATCC700603, *P. aeruginosa* ATCC27853, *E. faecalis* ATCC29212, *C. albicans* ATCC14053.

### Statistical analysis

The results of this study were analysed statistically using SYSTAT-13.2. Descriptive statistics using Microsoft Excel including frequencies, percentages, mean, median, and range were calculated for all demographic and microbiological variables. The results for antimicrobial susceptibility were expressed as proportions of resistant, sensitive, and intermediate isolates for each antibiotic per organism. Due to the small sample sizes in this study (OPD n = 13, IPD n = 31), with several antibiotic subgroups having tested denominators as low as 3, expected cell counts in the majority of 2×2 contingency tables fell below 5. Fisher’s Exact Test was therefore applied throughout as the primary statistical test for comparing resistance proportions between IPD and OPD settings, as it does not rely on large-sample normal approximation and remains valid regardless of cell size. A p-value of less than 0.05 was considered statistically significant. *Candida* spp. was excluded from ESBL, CRE, and MDR analyses as these mechanisms are not applicable to fungal organisms.

### Ethical considerations

This study was ethically approved by University Ethics Committee (Medical) of Swami Vivekanand Subharti University, Meerut Reference No: –SMC/UECM/2025/.11.6.8. Informed consent was waived because of the retrospective observational study design that involved analysis of existing anonymised laboratory data without direct patient contact. All data were handled in compliance with institutional confidentiality guidelines.

## RESULTS

### Characteristics of the patients

Majority of the patients were from the age of 61 – 75 years, followed by 46 – 60 age group. Both of these age groups accounted for 66 % of the total isolates. The average age calculated was 58 years. In terms of patient setting distribution, 74% of the isolates were from the IPD while 26% were from the OPD. The frequency of gender and patient setting distribution of UTI cases admitted to both IPD and OPD at Chhatrapati Shivaji Subharti Hospital from July 2025 – December 2025 is summarized in **Figure 1** below.

**Figure 1.**
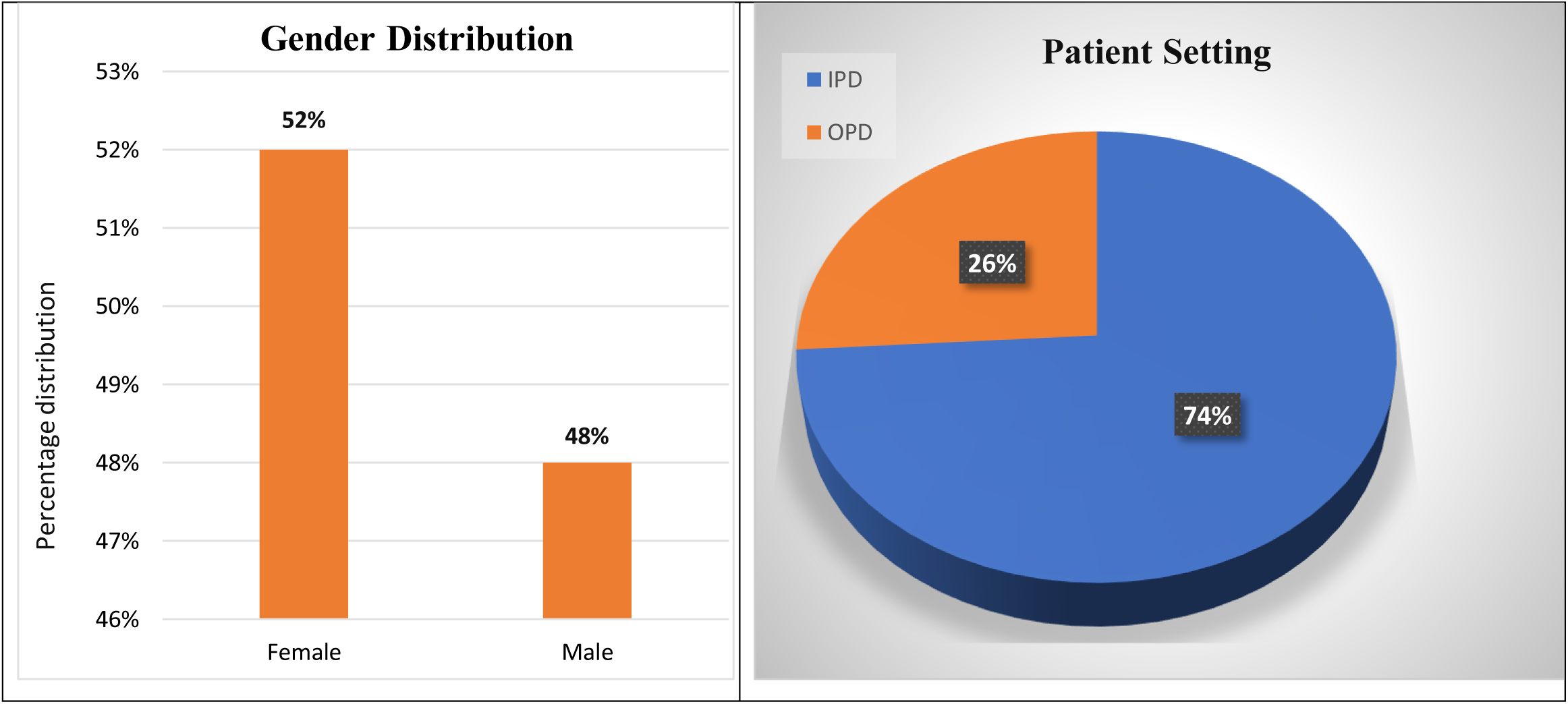
Gender and patient setting distribution. Females constituted 52% (n=26) of patients and males were 48% (n=24). Most uropathogens were isolated from IPD setting (IPD: n=37, 74%) while OPD accounted for 26% (n=13).

**Figure 2.**
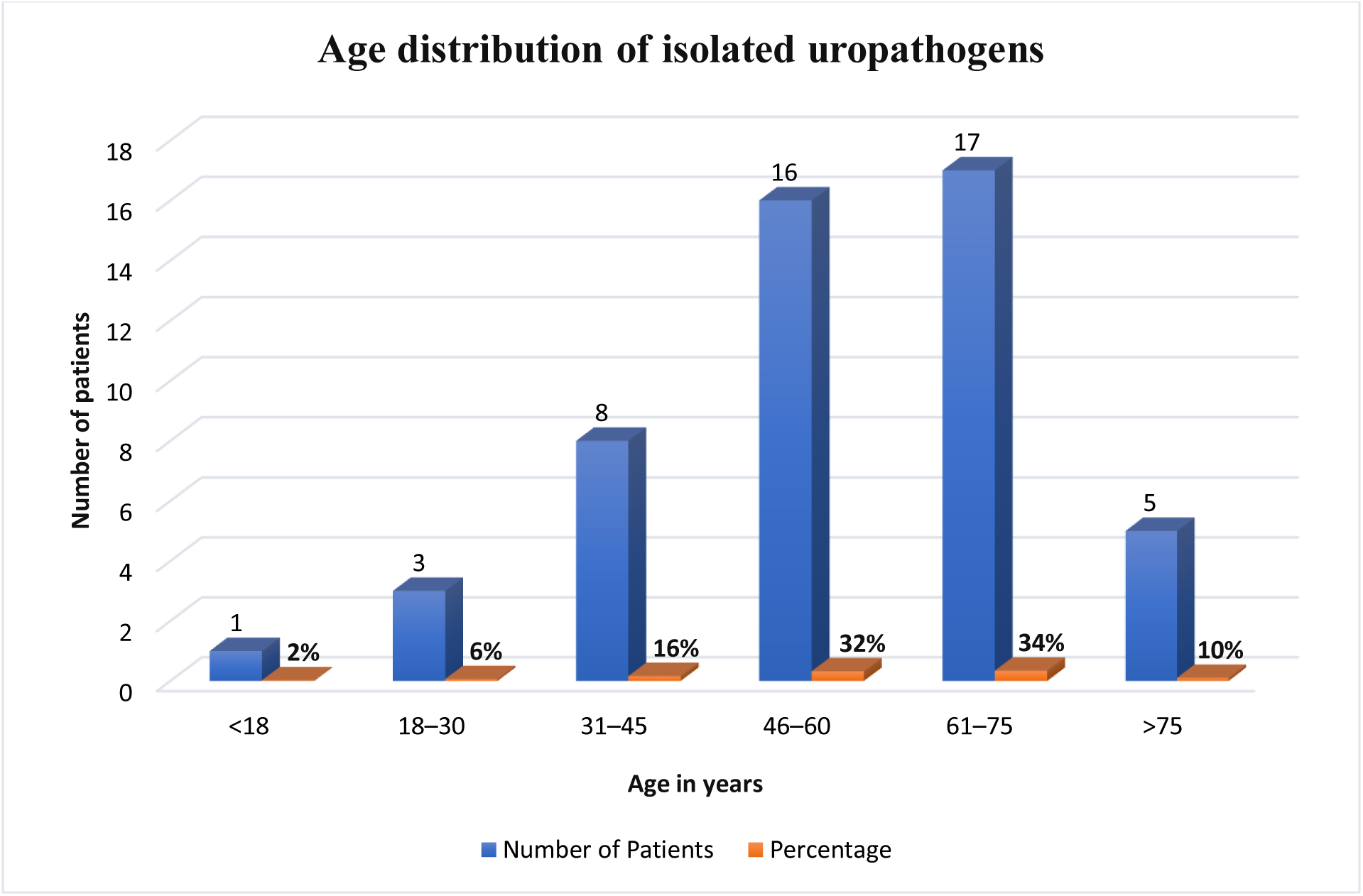
Age distribution of uropathogen isolates (N=50). The recorded age range was 18–95 years. The 61–75year age group recorded the highest number of isolates (n=17, 34%), followed by the 46–60year group (n=16, 32%). Mean age = 58.0 years.

### The Distribution of Pathogenic Bacteria

Intermediate results were excluded, Not Done (ND) isolates were not included in calculations, and p ≤ 0.05 was considered statistically significant. IPD had higher resistance rates than OPD. Commonly used antibiotics such ampicillin (92.0%), ceftriaxone (100%), ciprofloxacin (93.5%), cotrimoxazole (80.8%), piperacillin-tazobactam (89.3%), cefepime (96.4%), imipenem (82.1%), and meropenem (89.3%), had very high resistant rates especially in the IPD isolates. Nitrofurantoin (22.2% vs 68.2%, p = 0.0439) and meropenem (58.3% vs 89.3%, p = 0.0386), showed statically significant differences between IPD and OPD. Colistin (20% vs 8%) and fosfomycin (22.2% vs 39.1%) had lower resistance rates both in IPD and OPD.

### Antimicrobial Susceptibility Profiles of Major Uropathogens

**Figure 4** results show that *E. coli* had higher resistance to ceftriaxone (100%), ampicillin (100%), ciprofloxacin (96%), and cefepime (96%), and a higher sensitivity to fosfomycin (86%) and colistin (84%). In *K. pneumoniae,* extensive resistance to commonly used antibiotics including ceftriaxone, piperacillin-tazobactam, and amoxicillin-clavulanate (100% each) was observed; **Figure 5**. It showed susceptibility to colistin (83%) and fosfomycin (50%). In **Figure 6** *P. aeruginosa* showed complete resistance to ciprofloxacin (100%) and high resistance to amikacin, meropenem, imipenem, ceftazidime, and piperacillin-tazobactam (67% each), while colistin had the highest activity (67% sensitive). *E. faecalis,* a gram-positive isolate showed complete sensitivity to linezolid, teicoplanin, vancomycin, nitrofurantoin, and fosfomycin (100% each), but complete resistance to high-level gentamicin, tetracycline, and ciprofloxacin (100%); **Figure 7**.

**Figure 3:**
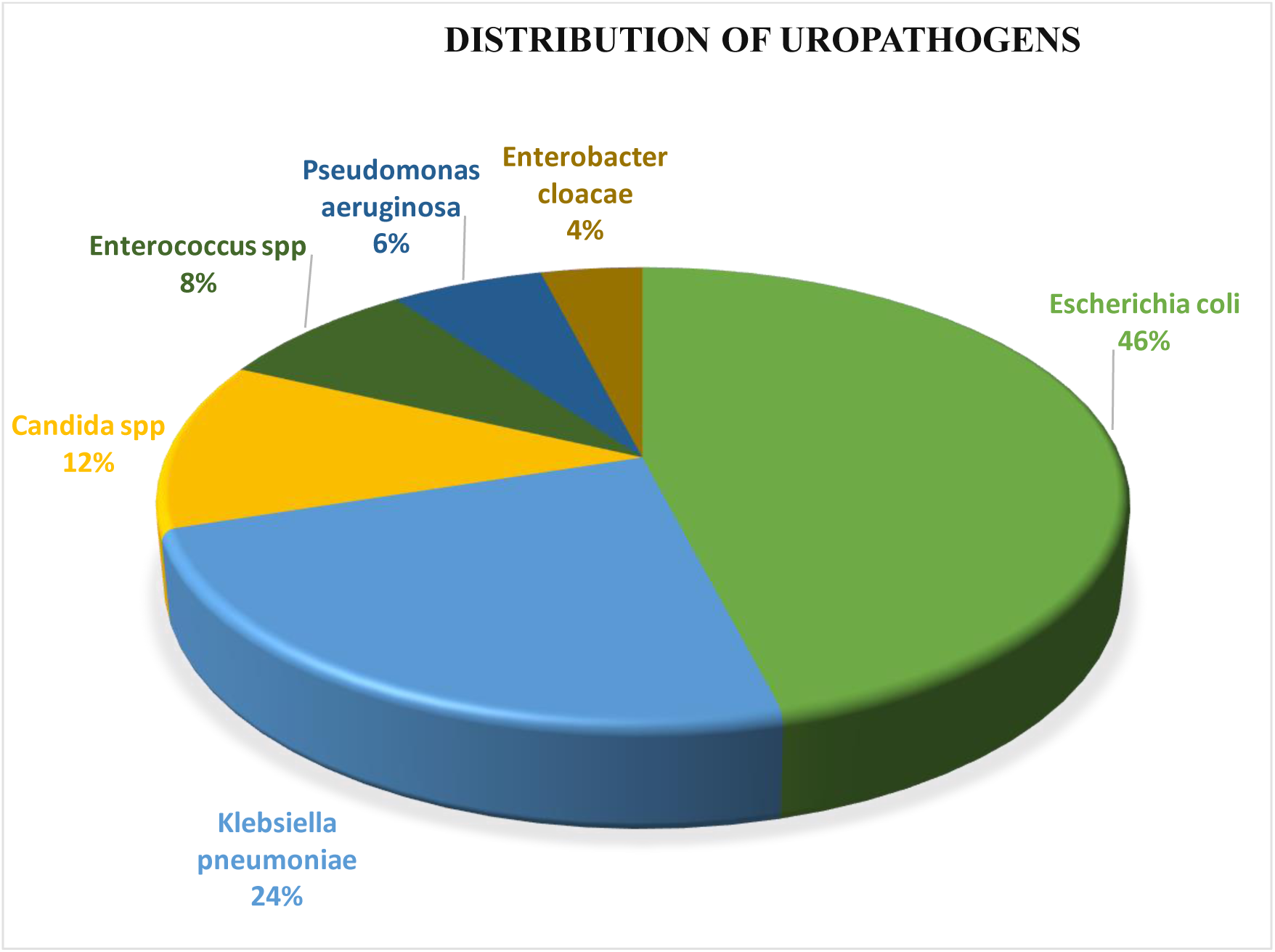
Distribution of Uropathogens. *Escherichia coli* (n=23, 46.0%), *Klebsiella pneumoniae* (n=12, 24.0%), *Candida* spp. (n=6, 12.0%), *Enterococcus* spp. (n=4, 8.0%), *Pseudomonas aeruginosa* (n=3, 6.0%), and *Enterobacter cloacae* (n=2, 4.0%). Gram-negative organisms collectively accounted for 40 of 50 isolates (80.0%), consistent *Candida* spp. accounted for 12% of isolates. The main pathogenic fungi isolated from the urine in the multi-speciality during July 2025 – December 2025 were *C. albicans*, *C. tropicalis,* and *C. glabrata*.

**Figure 4:**
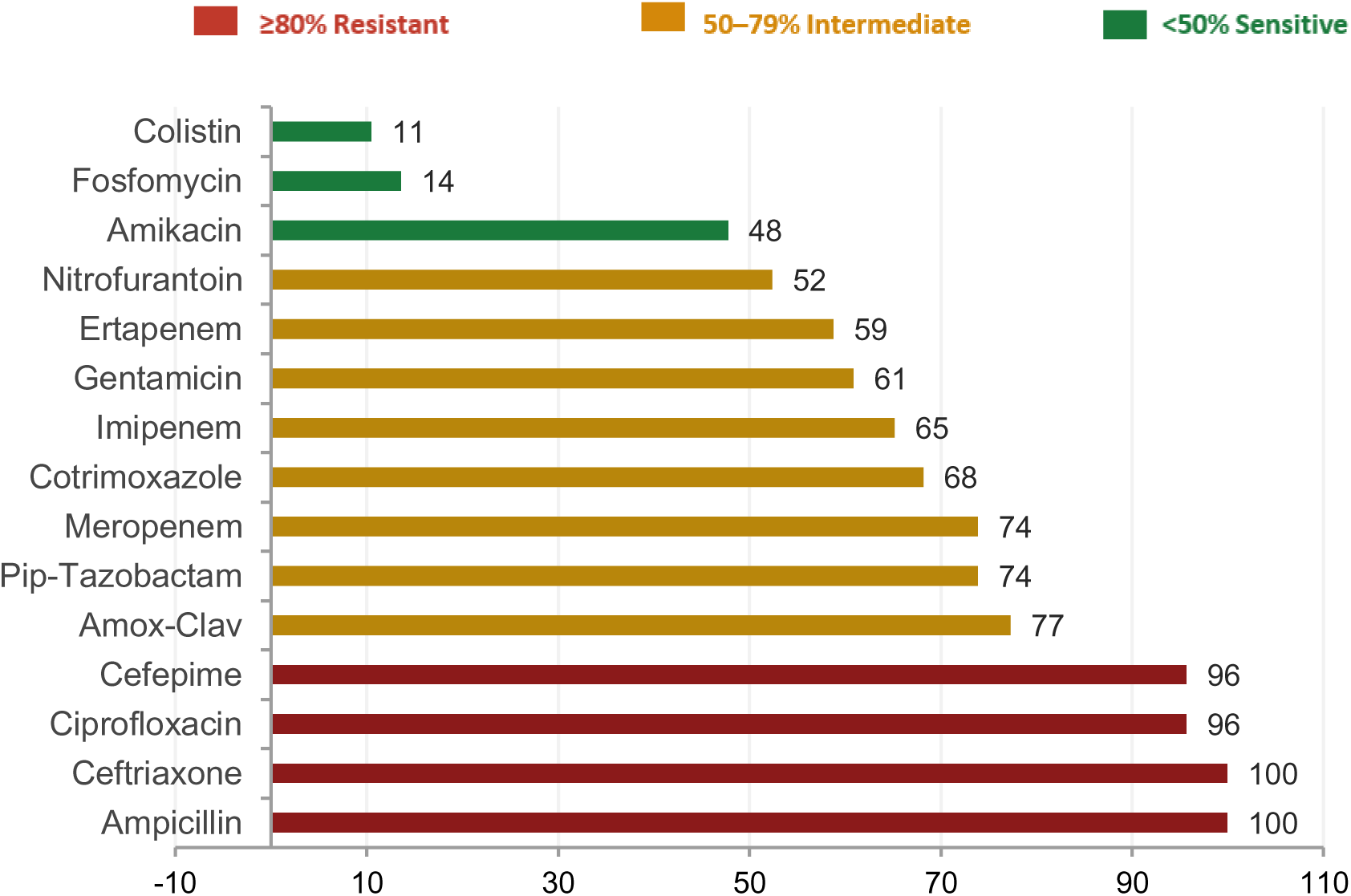
*Escherichia coli* Resistance Profile (n=23)

**Figure 5:**
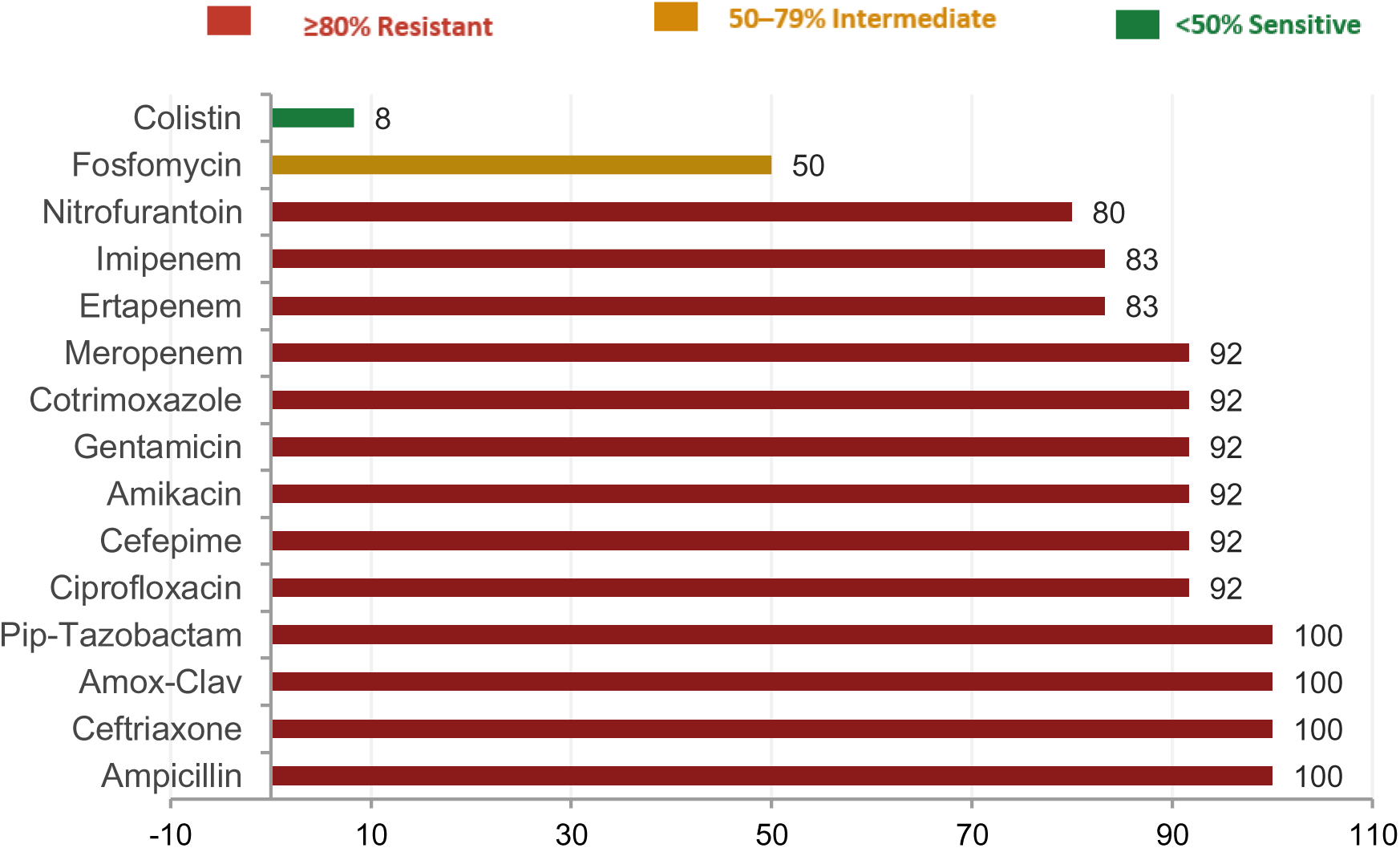
*Klebsiella pneumoniae* Resistance Profile (n=12)

**Figure 6:**
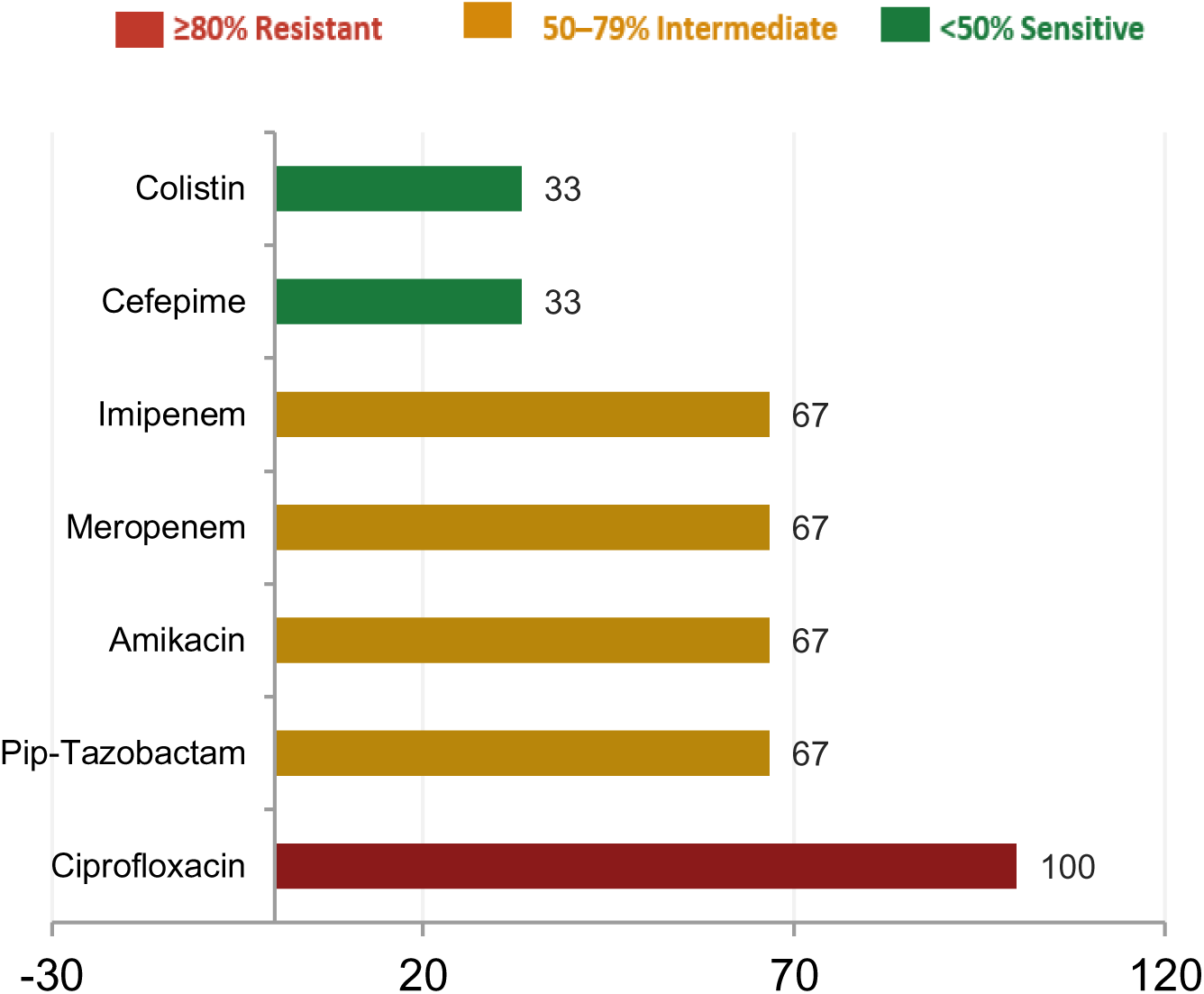
*Psuedomonas aeruginosa* (n=3) — Indicative Only.

**Figure 7:**
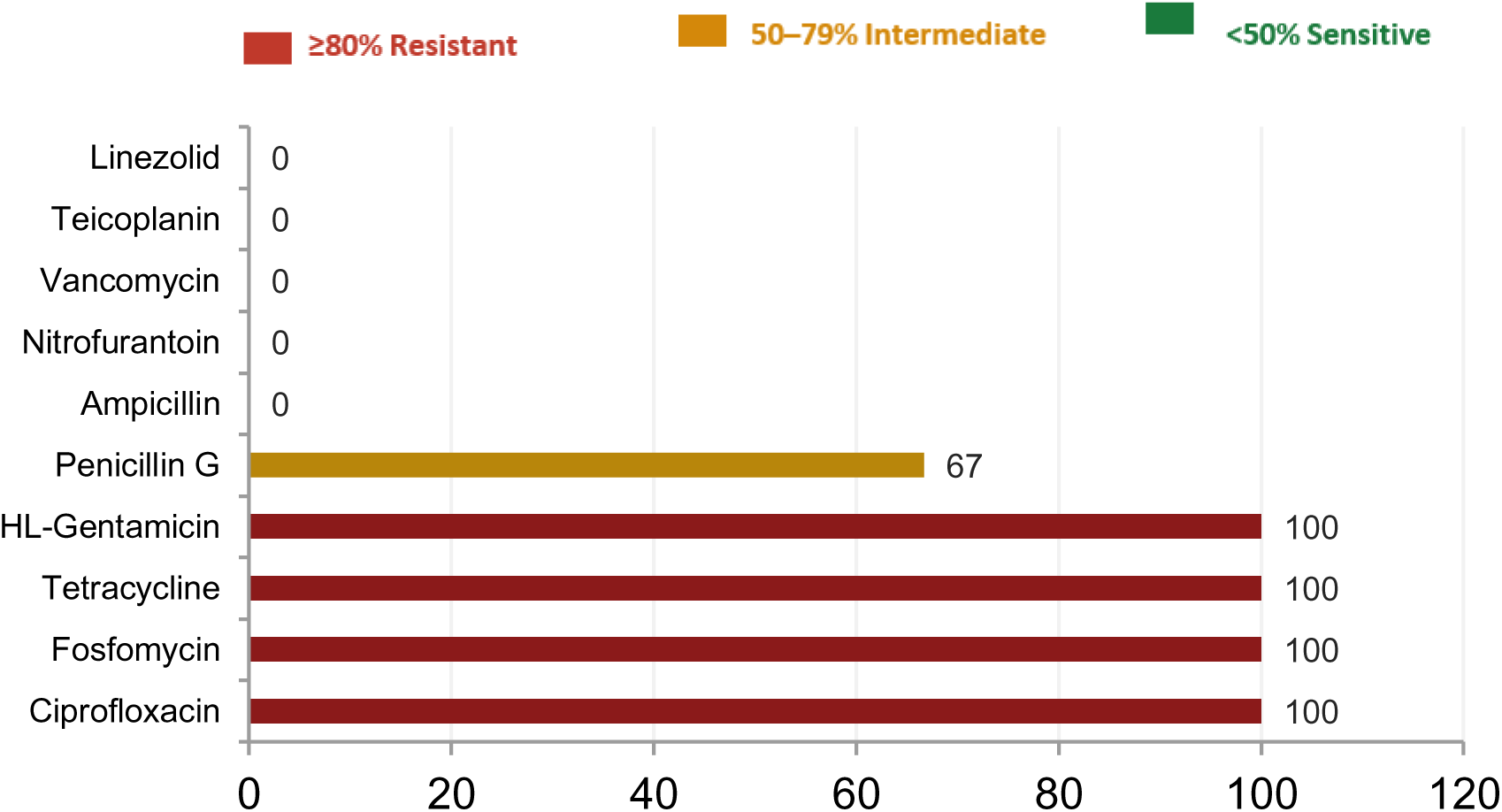
*Enterococcus faecalis* (n=3) — Gram-positive Pattern.

### Antifungal Susceptibility

**Table 4.2** is a representation of the antifungal susceptibility testing of the six *Candida* spp isolates that demonstrated complete susceptibility to all antifungal agents tested. All isolates were fully sensitive to fluconazole and voriconazole, while the five isolates tested against amphotericin B, caspofungin, and micafungin also showed complete sensitivity with no resistant or intermediate isolates detected. Species distribution included *Candida albicans* (n=3), *Candida tropicalis* (n=1), *Candida guilliermondii* (n=1), and *Candida lusitaniae* (n=1).

| Antibiotic | Tested (n) | Sensitive n (%) |
| --- | --- | --- |
| Fluconazole | 6 | 6 (100.0%) |
| Voriconazole | 6 | 6 (100.0%) |
| Amphotericin B | 5 | 5 (100.0%) |
| Caspofungin | 5 | 5 (100.0%) |
| Micafungin | 5 | 5 (100.0%) |

### ESBL, CRE and MDR Prevalence by Organism

**Table 4.3** Showing the ESBL, CRE and MDR prevalence by organism: Colour coding: Red >=80% | Amber 50-79% | Green <50% | Grey = N/A or not applicable per lab protocol. ESBL+CRE co-positivity (E. coli + K. pneumoniae): 31/35 tested = 88.6%. These isolates carry both ESBL and carbapenems simultaneously.

| Table 4.3 ESBL, CRE and MDR Prevalence by Organism |  |  |  |  |  |  |  |
| --- | --- | --- | --- | --- | --- | --- | --- |
| Organism | n | ESBL tested | ESBL+ n (%) | CRE tested | CRE+ n (%) | MDR n (%) | ESBL+CRE both+ |
| *E. coli* | 23 | 23 | 23 (100.0%) | 23 | 20 (87.0%) | 23 (100.0%) | 20 |
| *K. pneumoniae* | 12 | 12 | 12 (100.0%) | 12 | 11 (91.7%) | 12 (100.0%) | 11 |
| *Pseudomonas* | 3 | -- | N/A | 3 | 1 (33.3%) | 3 (100.0%) | N/A |
| *Enterococcus* | 4 | -- | N/A | -- | N/A | 4 (100.0%) | N/A |
| **Total bacterial** | **44** | 35 | 35 (100.0%) | 40 | 34 (85.0%) | 44 (100.0%) | 31 |

**Table 4.3** shows the prevalence of ESBL production, carbapenem resistance (CRE), and multidrug resistance (MDR) among uropathogens isolated in this study. All tested *E. coli* and *K. pneumoniae* isolates were found to be ESBL producers and multidrug resistant. Resistance to carbapenems was high with 87.0% of *E. coli* and 91.7% of *K. pneumoniae* isolates being carbapenem resistant. Most isolates were positive for both ESBL and CRE. All *Pseudomonas* isolates showed MDR and one isolate showed carbapenem resistance. Similarly, all *Enterococcus* isolates met MDR criteria.

## DISCUSSION

The present study describes the bacteriological profile and antimicrobial susceptibility patterns of 50 culture-positive urine isolates from patients attending Chhatrapati Shivaji Subharti Hospital, Meerut. Similar to the previous studies in Indian tertiary care centres and global epidemiological pattern of higher UTI incidence in women, more positive urine samples were found in females than in males as shown in **Figure 1** [5,11,13]. The close anatomical proximity of the anus to the reproductive organs and the lower parts of the urinary tract in females allows microorganisms like *Escherichia coli* and *Klebsiella pneumoniae* to opportunistically colonize the urinary system [14].

The results in **Figure 2** shows that the average age of the patients was 58 years and a median of 59.5 years. Overall, the most affected age group was between 61–75 years (n=17, 34.0%), followed by the 46–60 years group (n=16, 32.0%), together accounting for 66.0% of all isolates. These results are in line with the findings of the research done in Sichuan China [11]. Some of the risk factors of developing UTIs include previous history of UTIs, urinary catheterization, and conditions such as incontinence, all of which increase the susceptibility and recurrent infections [15].

### Inpatient (IPD) vs Outpatient (OPD)

Our results showed that IPD had a higher number of positive samples accounting for 37 of 50 isolates (74.0%) compared to 13 OPD isolates (26.0%) as depicted in **Figure 1**. Chooramani et al (2020) from Lucknow, North India, similarly found higher culture positivity rates in IPD compared to OPD patients [16]. Patients in intensive care units are more exposed to the main risk factors that increase their vulnerability to UTIs. These include indwelling urinary catheters, invasive procedures, prolonged antibiotic therapy which creates selective pressure, compromised immune status due to underlying illness, and comorbidities such as diabetes mellitus, hypertension, chronic kidney disease all of which may require prolonged bed rest. The emergency ICU had the highest burden with 14 of 50 isolates (28.0%), of which 7 were *E. coli* and 4 were *K. pneumoniae*. These results are similar to the findings of [17], who confirmed CAUTI prevalence of 73.75% in ICU patients, with patients over 60 years, hospitalised for more than 10 days, or with comorbidities such as diabetes and hypertension at highest risk. Their study also showed a demographic profile closely matching the present study’s patient population where the mean age was 58.0 years and IPD predominated.

Among all the antibiotics tested between IPD and OPD, the results in **Table 4.1** show that only nitrofurantoin (p = 0.044) and meropenem (p = 0.039) had a statistically significant difference. A higher resistance of carbapenem was observed in IPD isolates with meropenem resistance of 89.3% compared to 58.3% in OPD (p = 0.039). While imipenem resistance was higher in IPD isolates (82.1% vs 50.0%), this difference was not statistically significant (p = 0.056) likely due to the limited statistical power from the small sample size. Factors such as longer hospital stays, catheter use, and previous use of broad-spectrum antibiotics are likely to cause the marked difference in resistance of meropenem between IPD and OPD isolates [6,18,19,20].

Nitrofurantoin resistance was higher in IPD isolates (68.2% versus 22.2%, p = 0.044), indicating that the drug may still be useful for community-acquired UTIs but is less dependable for patients in hospital. Cefepime and ceftazidime also showed a higher resistance in the IPD isolates (96.4% vs 75.0%) and (100% vs 33.3%), but both did not reach statistical significance according to Fisher’s Exact Test (p = 0.073). This pattern is clinically meaningful and deserves further study in a larger sample. There was no statistical significance between settings with both fosfomycin and colistin leaving them as the most consistently effective antimicrobials for uropathogens across both clinical settings. Similarly, (Amladi et al., 2019 and Kalai et al., 2023) both identified fosfomycin and colistin as potential options for multidrug-resistant and carbapenem-resistant uropathogens, especially when they are used together with culture-based confirmation [21,22].

### Bacteriological Profile Distribution of Uropathogens

Of the 50 isolates, Gram-negative bacteria made up 40 (80.0%) of the total isolates, which aligns with global studies literature demonstrating that Gram-negative organisms, especially members of the *Enterobacteriaceae* family, are responsible for the overwhelming majority of UTIs in both community and hospital settings [23]. As shown in **Figure 3**, the main pathogenic bacteria isolated were *E. coli* (n=23, 46%), followed by *K. pneumoniae* (n=12, 24%). *Candida* spp. accounted for 12% of isolates (n=6). A similar finding was observed in global epidemiological reports and previous studies, where *E. coli* was the most common pathogen isolated demonstrating this pathogen is the most common cause of both community-acquired and healthcare-associated UTIs [5,24,25]. It is suggested that both *E. coli* and *K. pneumoniae* are the primary drivers of UTI burden as they constituted 70% of all uropathogens which aligns with the study done by [26].

The proportion of *Klebsiella pneumoniae* observed in the present study (24.0%) was higher than that reported in several studies, where its prevalence typically ranges between 10–20% [23,26,27,28]. It also shows the IPD predominant patient population of the present study in which catheter-associated infection and prolonged antibiotic exposure create conditions particularly favourable for *K. pneumoniae* acquisition and persistence.

While most research studies report only *C. albicans* or *C. tropicalis*, our study showed four different *Candida* species namely *C. albicans*, *C. tropicalis*, *C. guilliermondii*, and *C. lusitaniae. C. guilliermondii* and *C. lusitaniae* are rare fungi that are infrequently reported in Indian uropathogen surveillance literature. The presence of these rare species in ICU patients warrants clinical vigilance beyond standard antifungal coverage [29]. Non-albicans *Candida* species (NAC) were greater in number (4/6, 66.7%) which reflects the worldwide reported epidemiological shift towards NAC in healthcare settings [30]. Although *Proteus mirabilis*, *Acinetobacter*, and *Staphylococcus aureus* were missing from this study, these three organisms commonly appear in both international and Indian studies [16,23,24,27].

### Antimicrobial Resistance Profiles of Major Uropathogens

*Escherichia coli* had complete resistance to ampicillin 100% (23/23) and ceftriaxone 100% (18/18), and very high resistance to ciprofloxacin and cefepime at 95.7% (22/23). These findings align with a five-year retrospective study from Romania that showed a high ampicillin resistance of 48–55% in *E. coli* and concluded that these agents should be avoided for empirical UTI treatment. A Saudi Arabian tertiary care hospital study also reported cephalexin resistance of 95.45% and ampicillin resistance of 88.46% [31,32].

*E. coli* also showed a substantial resistance to carbapenems with meropenem 73.9% (17/23), imipenem 65.2% (15/23), and ertapenem 58.8% (10/17), which shows that there is a rise in the carbapenem-resistant strains. Additionally, treatment options have further been limited due to resistance to commonly used antimicrobials such as amoxicillin-clavulanate 77.3% (17/22), piperacillin-tazobactam 73.9% (17/23), gentamicin 60.9% (14/23), and cotrimoxazole 68.2% (15/22). While nitrofurantoin resistance was moderate 52.4% (11/21), fosfomycin 86.4% (19/22), and colistin 84.2% (19/22), had high sensitivity. Similarly, a study conducted in a tertiary care hospital in India, Jaipur found a high level of carbapenem resistance, with most isolates producing carbapenemase enzymes, predominantly the blaOXA-48 type. This enzymatic resistance mechanism may explain the high level of carbapenem resistance observed in the present study [33]. Therefore, with carbapenems failing in nearly three-quarters of *E. coli* isolates, treatment with carbapenem which has traditionally been the last reliable option is no longer justified at this institution without prior susceptibility confirmation.

*Klebsiella pneumoniae* showed a much higher resistance burden with 100% resistance to amoxicillin-clavulanate, ceftriaxone, piperacillin-tazobactam, and ampicillin, **Figure 5**. A high resistance was also seen for ciprofloxacin, cefepime, amikacin, gentamicin, and cotrimoxazole 91.7% (11/12). Moreover, carbapenem resistance was near-total, with meropenem 91.7% (11/12) and both ertapenem and imipenem 83.3% (10/12), leaving no reliable β-lactam options. In comparison to *E. coli*, *K. pneumoniae* showed a higher resistance with all the key antimicrobials including cotrimoxazole, aminoglycosides, and carbapenems. Colistin remained the only effective agent with sensitivity of 83.3% (10/12), while fosfomycin showed limited activity 50.0% (2/4). Similar to earlier studies, the results show that the observed high level of resistance to β-lactam antibiotics indicates a limited effectiveness of the traditionally used antimicrobials [34].

Despite being limited by a small sample size, *Pseudomonas aeruginosa* had a worrying resistance profile with 100% resistance to ciprofloxacin (3/3) and 66.7% (2/3) resistance to piperacillin-tazobactam, amikacin, meropenem, and ceftazidime; **Figure 6**. Colistin is the main antibiotic used for treatment of these infections since it maintained sensitivity in 66.7% (2/3) of isolates. Since the sample size was only 3, the results are not reliable enough to make definitive conclusions and should be confirmed with a larger study.

*Enterococcus faecalis* had Gram-positive resistance pattern with a complete resistance observed to ciprofloxacin, fosfomycin, tetracycline 100% (3/3) and high-level gentamicin 100% (2/2) as shown in **Figure 7**. However, full sensitivity was retained for ampicillin 100% (3/3), nitrofurantoin 100% (3/3), vancomycin 100% (2/2), teicoplanin 100% (3/3), and linezolid 100% (3/3). The resistance pattern observed for *Enterococcus faecalis* in the present study is largely consistent with previous findings, showing high resistance to fluoroquinolones, tetracyclines, and aminoglycosides, while retaining susceptibility to linezolid and glycopeptides. However, the detection of vancomycin moderate resistance in this study suggests the possible emergence of resistant strains, warranting continued surveillance [16].

All the isolated *Candida spp* had a 100% sensitivity to antifungal agents, such as fluconazole, voriconazole, amphotericin B, caspofungin, and micafungin, indicating preserved antifungal efficacy in this setting; **Table 4.2**. These findings are consistent with studies reporting preserved antifungal efficacy in certain clinical settings. However, the continued rise in antifungal resistance and the emergence of non-*albicans Candida* species underscores the importance of continued surveillance and antifungal stewardship [12,35].

### ESBL, CRE, and MDR Burden

The study revealed interesting ESBL, CRE and MDR results in which all the tested isolates were 100% ESBL-positive, 85.0% of tested Gram-negative isolates (34/40) were CRE-positive; **Table 4.3.** 100% of non-fungal bacterial isolates met MDR criteria. Both *E. coli* and *K. pneumoniae* demonstrated 100% ESBL positivity, with CRE rates of 87.0% and 91.7% respectively, ESBL and CRE co-positivity was identified in 31 of 35 tested *Enterobacteriaceae* isolates (88.6%). These results are above the 44.8% ESBL prevalence findings by [26] in community-acquired UTI, and the 39.1% reported by Asokan et al (2025) [36]. The CRE positivity was 85.0% of Gram-negative isolates tested which was also above the 4.3% carbapenem resistance reported by [26] in community OPD isolates. Similarly, a high number of Gram-negative bacteria were carbapenem-resistant, with *Klebsiella pneumoniae* showing the greatest resistance burden [37].

This is consistent with a decade-long retrospective study from a South Indian tertiary care hospital which documented carbapenem resistance in *K. pneumoniae* rising from 3.70% in 2014 to a peak of 66.13% in 2021, reflecting the particularly severe resistance environment of an ICU-predominant North Indian tertiary care setting [38]. This confirms that the resistance burden at this hospital is of a wholly different order of magnitude compared to community-level data. MDR was universal at 100% of all 44 bacterial isolates, which highlights the complete failure of empirical treatment with common antibiotic classes; Table 4.3. A 2025 Scientific Reports study on carbapenem-resistant and ESBL-producing MDR *E. coli* from UTI patients found that 66.26% of isolates were MDR, a figure already considered alarming at publication but substantially lower than the 100% MDR rate in the present study [39].

Most of the bacteria simultaneously carried both resistance mechanisms as reflected by the co-positivity of ESBL and CRE in the tested *Enterobacteriaceae* isolates. Harsha and Kashid (2026) mention that the simultaneous production of ESBL and carbapenemase enzymes among uropathogenic *E. coli* is becoming more common in Indian tertiary care hospitals, creating a dual-resistance phenotype that reduces treatment options [40].

Interestingly, the results show that resistance burden is not only in hospital environments. This is reflected by OPD isolates which demonstrated 100% ESBL positivity (11/11 tested) and 100% MDR (13/13), with a CRE rate of 75.0% (9/12 tested), indicating that carbapenem-resistant, ESBL-producing, multidrug-resistant uropathogens are already circulating in the community population attending this hospital. With New Delhi metallo-beta-lactamase being the most common carbapenemase mechanism among *E. coli* and *K. pneumoniae* in both inpatient and outpatient isolates, a study on carbapenem resistance trends using the same VITEK-2 Compact system as the current study confirmed that carbapenem resistance is endemic in the Indian subcontinent. This suggests that the community CRE burden in the Meerut region reflects the established circulation of NDM-producing strains consistent with patterns documented across North India [18].

Overall, the study shows extremely high levels of antibiotic resistance, including widespread ESBL, carbapenem resistance, and multidrug resistance, even at the community level. The reduced effectiveness of commonly used antibiotics, mostly β-lactams and fluoroquinolones, highlights the need for routine culture and sensitivity testing and the importance of antimicrobial stewardship. The retained activity of fosfomycin for *E. coli* and colistin for highly resistant Gram-negative organisms suggests their role as key therapeutic options, particularly in tertiary care hospitals such as in Meerut. These findings highlight the urgent need to introduce a formal antibiotic stewardship program at Chhatrapati Subharti Hospital, supported by regularly updated local antibiograms. As the first baseline study from this hospital, the results are important for guiding antibiotic use, improving infection control, and strengthening public health monitoring in Meerut and surrounding regions.

## CONCLUSION

This study provided a microbiological audit of culture positive uropathogens and their profiles of antibiotic resistance at Chhatrapati Shivaji Subharti Hospital, Meerut. Of the uropathogens isolated, *E. coli* and *K. pneumoniae* accounted for 70% of the isolates with four *Candida* species that included rarely reported *C. guilliermondii* and *C. lusitaniae* among the fungal uropathogens. Most of the uropathogens were isolated from IPD patients showing a much higher UTI burden in critically ill, catheterised, and comorbid patients. These findings indicate a very severe pattern of resistance in line with the increasing frequency of combined ESBL and carbapenems producing organisms in Indian tertiary care hospitals. These results also show that the use of commonly used antibiotics such as ampicillin, amoxicillin-clavulanate, and all cephalosporin should be discontinued within this hospital.

One of the most important findings was the detection of complete ESBL positivity and MDR among OPD isolates, with a high rate of CRE resistance, demonstrating that this resistance burden is not only in the hospital environment but is also circulating in the community population of the Meerut region. Conversely, all *Candida* isolates were sensitive to antifungal drugs, which is reassuring; however, the presence of different and rare species highlights the need for proper species identification in future studies.

In conclusion, the results of this study demonstrate that fosfomycin which is sensitive in *E. coli* and colistin which is sensitive in *K. pneumoniae* are the only remaining agents with clinically useful activity against the main Gram-negative uropathogens. Their protection through antimicrobial stewardship is a matter of clinical urgency. This study also provides the crucial baseline microbiological information, the local institutional antibiogram upon which evidence-based empirical prescribing guidelines, infection control policies, and antibiotic stewardship programmes must be developed at Chhatrapati Subharti Hospital.

## STUDY LIMITATIONS

The main limitation of this study was the small sample size, especially the limited number of culture-positive OPD isolates (n=13). This made it difficult to perform strong statistical comparisons between OPD and IPD isolates for most antibiotics. Other limitations include the retrospective single-centre design, the absence of molecular confirmation of resistance genes (bla-NDM, bla-CTX-M, bla-OXA-48), and the use of disk diffusion rather than MIC-based testing for *Candida* susceptibility, which may not detect reduced susceptibility near resistance breakpoints for rare species such as *C. guilliermondii* and *C. lusitaniae*.

In future, studies should focus on molecular characterisation of resistance genes including *bla*-NDM, *bla*-OXA-48, *bla*-KPC, and *bla*-CTX-M to better understand how resistant bacteria spread between patients and hospital wards. In addition to that longitudinal surveillance of at least 12–24 months is necessary to identify emerging resistance to fosfomycin and colistin before these last-resort agents are lost clinically. Larger prospective studies should divide isolates by ICU, general ward, and OPD while also considering additional factors such as catheter use, prior antibiotic exposure, and comorbidities such as diabetes.

More accurate antifungal testing methods, such as the MIC (Minimum Inhibitory Concentration) testing, should be used for rare *Candida* species such as *Candida guilliermondii* and *Candida lusitaniae*. This is because disk diffusion testing may not always accurately show whether these fungi are resistant or sensitive to antifungal drugs, especially when the results are close to the resistance cut-off point.

Finally, future research should investigate whether using the institutional antibiogram helps prescribe antibiotics more appropriately and improves patient outcomes. Research should also connect hospital and community surveillance systems using a One Health approach, which recognises that antimicrobial resistance is linked to human, animal, and environmental health, in line with India’s National Action Plan on Antimicrobial Resistance.

## THE USE OF AI

During the preparation of this research article, the authors used Claude to assist with preparation of figures and tables. After using this tool, the authors reviewed and edited the content as needed and take full responsibility for the content of this publication.

## Supporting information

file:///C:/Users/HP/AppData/Local/Temp/075c415c-7a58-4ae7-a702-e8e128e7eec8_SUPPLEMENTARY%20MATERIAL%20EDITED%20PDF%20ZIP.zip.ec8/SUPPLEMENTARY%20MATE

## Data Availability

All data produced in the present study are available upon reasonable request to the authors
All data produced in the present work are contained in the manuscript

## REFERENCES

1. Zeng Z, Zhan J, Zhang K, Chen H, Cheng S. Global, Regional, and National Burden of Urinary Tract Infections from 1990-2019: an Analysis of the Global Burden of Disease Study 2019 [Internet]. In Review; 2021 [cited 2026 Feb 26]. Available from: https://www.researchsquare.com/article/rs-829349/v1 doi: 10.1007/s00345-021-03913-0

2. Yang X, Chen H, Zheng Y, Qu S, Wang H, Yi F. Disease burden and long-term trends of urinary tract infections: A worldwide report. Front Public Health. 2022 Jul 27;10:888205. doi:10.3389/fpubh.2022.888205

3. Gupta A, Sharma B. Recent trends in uropathogenic infections in patients of a tertiary care center, New Delhi, India, – a topic of urgent attention. Microbiology Independent Research Journal (MIR Journal). 2023;10(1). doi:10.18527/2500-2236-2023-10-1-39-44

4. He Y, Zhao J, Wang L, Han C, Yan R, Zhu P, et al. Epidemiological trends and predictions of urinary tract infections in the global burden of disease study 2021. Sci Rep. 2025 Feb 8;15(1):4702. doi: 10.1038/s41598-025-89240-5

5. Sharma R, Parihar S, Kinimi SV, Choudhary S. An Observational Study from Northern India to Evaluate Catheter-associated Urinary Tract Infection in Medical Intensive Care Unit at a Tertiary Care Centre. Indian Journal of Critical Care Medicine. 2023 Aug 31;27(9):642–6. doi: 10.5005/jp-journals-10071-24519

6. Akhavizadegan H, Hosamirudsar H, Pirroti H, Akbarpour S. Antibiotic resistance: a comparison between inpatient and outpatient uropathogens. East Mediterr Health J. 2021 Feb 1;27(2):124–30. doi: 10.26719/emhj.20.085

7. Saini V, Longkumer I, Malik S, Kumari A. Antibiotic Resistance Trends in Urinary Tract Infections among Women Receiving Obstetric and Gynaecological Care: A Two-Year Study from India. J Pure Appl Microbiol. 2026 Mar 1;20(1):209. doi:10.22207/JPAM.20.1.06

8. Gharavi MJ, Zarei J, Roshani-Asl P, Yazdanyar Z, Sharif M, Rashidi N. Comprehensive study of antimicrobial susceptibility pattern and extended spectrum beta-lactamase (ESBL) prevalence in bacteria isolated from urine samples. Sci Rep. 2021 Jan 12;11(1):578. doi: 10.1038/s41598-020-79791-0

9. Ahmed SK, Hussein S, Qurbani K, Ibrahim RH, Fareeq A, Mahmood KA, et al. Antimicrobial resistance: Impacts, challenges, and future prospects. Journal of Medicine, Surgery, and Public Health. 2024 Apr;2:100081. doi:10.1016/j.glmedi.2024.100081

10. Sharma A, Thakur N, Thakur A, Chauhan A, Babrah H. The Challenge of Antimicrobial Resistance in the Indian Healthcare System. Cureus. 2023 Jul 21. doi: 10.7759/cureus.42231

11. Xu X, Wang Y, Li N, Jin Y, Xu X, Zhou Z, et al. Uropathogen profiles and their antimicrobial resistance patterns in patients: a three-year retrospective study in Sichuan region. Front Public Health. 2025 Feb 27;13:1493980. doi:10.3389/fpubh.2025.1493980

12. Woldemariam HK, Geleta DA, Tulu KD, Aber NA, Legese MH, Fenta GM, et al. Common uropathogens and their antibiotic susceptibility pattern among diabetic patients. BMC Infect Dis. 2019 Dec;19(1):43. doi: 10.1186/s12879-018-3669-5

13. Priyadarshini A. Antimicrobial Resistance Pattern and Uropathogen Distribution of Pediatric UTIin Vadodara. AJBR. 2024 Nov 1;5835–40. doi:10.53555/AJBR.v27i3S.3438

14. Baimakhanova B, Sadanov A, Trenozhnikova L, Balgimbaeva A, Baimakhanova G, Orasymbet S, et al. Understanding the Burden and Management of Urinary Tract Infections in Women. Diseases. 2025 Feb 15;13(2):59. doi:10.3390/diseases13020059

15. Rodriguez-Mañas L. Urinary tract infections in the elderly: a review of disease characteristics and current treatment options. DIC. 2020 Jul 8;9:1–8. doi: 10.7573/dic.2020-4-13

16. Chooramani G, Jain B, Chauhan PS. Prevalence and antimicrobial sensitivity pattern of bacteria causing urinary tract infection; study of a tertiary care hospital in North India. Clinical Epidemiology and Global Health. 2020 Sep;8(3):890–3. doi:10.1016/j.cegh.2020.02.018

17. Purandare B, Jha T, Khaparde M, Parkhe TS, Lavate R. Incidence Risk Factors and Drug Resistance Patterns of Bacterial Isolates in Patients with Catheter-associated Urinary Tract Infections. Indian Journal of Critical Care Medicine. 2025 Mar 31;29(4):338–44. doi: 10.5005/jp-journals-10071-24932

18. Chatterjee N, Nirwan PK, Srivastava S, Rati R, Sharma L, Sharma P, et al. Trends in carbapenem resistance in Pre-COVID and COVID times in a tertiary care hospital in North India. Ann Clin Microbiol Antimicrob. 2023 Jan 3;22(1):1. doi: 10.1186/s12941-022-00549-9

19. Debbarma M, Behera B, Rout B, Panigrahy R, Baral P. A Glimpse into the Resistant Pattern of Uropathogens: An Overview. J Pure Appl Microbiol. 2022 Dec 1;16(4):2310–6. doi:10.22207/JPAM.16.4.35

20. Eshwarappa M, Gangula RS, Rajashekar R, Prabhu PP, Hamsa V, Yousuff M, et al. Clinical, Microbiological Profile, and Treatment Outcomes of Carbapenem-Resistant Urinary Tract Infections in a Tertiary Care Hospital. Indian J Nephrol. 2024 Jul 15;35:53–8. doi: 10.25259/ijn_530_23

21. Amladi AU, Abirami B, Devi SM, Sudarsanam TD, Kandasamy S, Kekre N, et al. Susceptibility profile, resistance mechanisms & efficacy ratios of fosfomycin, nitrofurantoin & colistin for carbapenem-resistant Enterobacteriaceae causing urinary tract infections. Indian Journal of Medical Research. 2019 Feb;149(2):185–91. doi: 10.4103/ijmr.IJMR_2086_17

22. Kalai J, Maheswary D, Leela KV, Gopinathan A. Susceptibility Profile of Nitrofurantoin and Fosfomycin among Carbapenem-resistant Enterobacteriaceae Isolates in UTI from a Tertiary Care Hospital. J Pure Appl Microbiol. 2023 Mar 5;17(1):345–53. doi:10.22207/JPAM.17.1.24

23. Flores-Mireles AL, Walker JN, Caparon M, Hultgren SJ. Urinary tract infections: epidemiology, mechanisms of infection and treatment options. Nat Rev Microbiol. 2015 May;13(5):269–84. doi: 10.1038/nrmicro3432

24. Khan S, Maroof P, Amin U. Microbial Etiology and Resistance Patterns of Urinary Tract Infection at a Tertiary Care Centre – A Hospital based Study. J Pure Appl Microbiol. 2023 Sep 1;17(3):1659–68. doi:10.22207/JPAM.17.3.28

25. Wu W, Wu Q, Zhong W, Han L. Distribution and Drug-Resistance Analysis of Uropathogens in Urinary Tract Infections. Khurshid M, editor. Canadian Journal of Infectious Diseases and Medical Microbiology. 2026 Jan;2026(1):1700474. doi: 10.1155/cjid/1700474

26. Mohapatra S, Panigrahy R, Tak V, J. V. S, K. C. S, Chaudhuri S, et al. Prevalence and resistance pattern of uropathogens from community settings of different regions: an experience from India. Access Microbiology. 2022 Feb 28;4(2). doi: 10.1099/acmi.0.000321

27. Gaur S, Gahlot R, Netam S, Dadarya S, Sherwani N. Epidemiology, microbiological profile and susceptibility pattern of uropathogens from a tertiary care hospital in central India. IJMR. 2024 Dec 28;11(4):277–82. doi:10.18231/j.ijmr.2024.048

28. Odoki M, Almustapha Aliero A, Tibyangye J, Nyabayo Maniga J, Wampande E, Drago Kato C, et al. Prevalence of Bacterial Urinary Tract Infections and Associated Factors among Patients Attending Hospitals in Bushenyi District, Uganda. International Journal of Microbiology. 2019 Feb 17;2019:1–8. doi: 10.1155/2019/4246780

29. Kande S, Patro S, Panigrahi A, Khora P, Pattnaik D. Prevalence of uropathogens and their antimicrobial resistance pattern among adult diabetic patients. Indian J Public Health. 2021;65(3):280. doi: 10.4103/ijph.IJPH_1413_20

30. Kullberg BJ, Arendrup MC. Invasive Candidiasis. Campion EW, editor. N Engl J Med. 2015 Oct 8;373(15):1445–56. doi: 10.1056/NEJMra1315399

31. Alzahrani M, Ali M, Anwar S. Bacteria causing urinary tract infections and its antibiotic susceptibility pattern at tertiary hospital in Al-Baha region, Saudi Arabia: A retrospective study. J Pharm Bioall Sci. 2020;12(4):449. doi: 10.4103/jpbs.JPBS_294_19

32. Goleanu (Vasiloiu) CD, Vrancianu CO, Goleanu DA, Tantu MM, Csutak O. Trends in Antibiotic Resistance of Escherichia coli Strains Isolated from Clinical Samples (2019– 2023): A Hospital-Based Retrospective Analysis. Pathogens. 2025 Sep 13;14(9):927. doi: 10.3390/pathogens14090927

33. Rajni E, Bairwa K, Galav H, Upadhyaya H, Gajjar D. An update on carbapenem-resistant Enterobacterales: A prospective study from Western India. Journal of Postgraduate Medicine. 2025 Apr;71(2):61–7. doi: 10.4103/jpgm.jpgm_558_24

34. Bhargava K, Nath G, Bhargava A, Kumari R, Aseri GK, Jain N. Bacterial profile and antibiotic susceptibility pattern of uropathogens causing urinary tract infection in the eastern part of Northern India. Front Microbiol. 2022 Aug 9;13:965053. doi: 10.3389/fmicb.2022.965053

35. Mishra N, Kumari D, Mishra A. Prevalence of Candida species in Urinary Tract Infections from a Tertiary Care Hospital at Lucknow, Uttar Pradesh, India: A Retrospective Study. NJLM. 2022. doi: 10.7860/NJLM/2022/56319.2676

36. Asokan S, Jacob T, Jacob J, AlSosowaa AA, Vijayan S. Trends in antimicrobial susceptibility patterns of Klebsiella pneumoniae isolated from clinical samples at a tertiary care hospital in Kerala, India. Next Research. 2025 Sep;2(3):100654. doi: 10.1016/j.nexres.2025.100654

37. Sahu S, Patil P, Ray S, Aditi. Continued Efficacy Of Nitrofurantoin And Co-Trimoxazole For Community-Acquired Urinary Tract Infections Amid Rising Antimicrobial Resistance In Southern India. Vol. 11. 2025;11. doi: 10.64252/2es1vg55

38. Neelambike Sumana M, Maheshwarappa YD, G. K, Mahale RP, G. S S, Raghavendra Rao M, et al. A retrospective study of the antimicrobial susceptibility patterns of Klebsiella pneumoniae isolated from urine samples over a decade in South India. Front Microbiol. 2025 Jun 17;16:1553943. doi:10.3389/fmicb.2025.1553943

39. Sivarajan V, Ganesh AV, Subramani P, Ganesapandi P, Sivanandan RN, Prakash S, et al. Prevalence and genomic insights of carbapenem resistant and ESBL producing Multidrug resistant Escherichia coli in urinary tract infections. Sci Rep. 2025 Jan 20;15(1):2541. doi: 10.1038/s41598-024-84754-w

40. Harsha TK, Kashid RA. Identification of ESBL and Carbapenemase Genes and their Correlation with Antibiotic Resistance Profiles in Escherichia coli Strains Isolated from Patients with Urinary Tract Infections in a Tertiary Care Hospital. J Pure Appl Microbiol. 2026 Mar 1;20(1):488. doi: 10.22207/JPAM.20.1.35

