## Supplementary material for "Bacteriological Profile and Antimicrobial Susceptibility Patterns of Uropathogens at a Tertiary Care Hospital in North India: A Retrospective Descriptive Study": file:///C:/Users/HP/AppData/Local/Temp/075c415c-7a58-4ae7-a702-e8e128e7eec8_SUPPLEMENTARY%20MATERIAL%20EDITED%20PDF%20ZIP.zip.ec8/SUPPLEMENTARY%20MATE: SUPPLEMENTARY MATERIAL EDITED PDF.pdf

### MASTER CHART

| S.No | Patient ID | Age Range (Yrs) | Sex (M/F) | Ward / Setting | OPD/IPD | Collection Month/Year | Sample Type | Organism Isolated | Colony Count (CFU/mL) | Gram Stain | Identification Method | Lab No. |
|---|---|---|---|---|---|---|---|---|---|---|---|---|
| 1 | PT-001 | 55-59 | F | EMERGENCY ICU | IPD | Dec 2025 | Urine (MSU) | Escherichia coli | ≥10^5 | Gram-negative | Automated ID & Sensitivity | 6681 |
| 2 | PT-002 | 70-74 | M | MEDICINE WARD-I | IPD | Dec 2025 | Urine (MSU) | Pseudomonas aeruginosa | ≥10^5 | Gram-negative | Automated ID & Sensitivity | 6491 |
| 3 | PT-003 | 60-64 | F | RESPIRATORY ICU | IPD | Dec 2025 | Urine (MSU) | Escherichia coli | ≥10^5 | Gram-negative | Automated ID & Sensitivity | 6344 |
| 4 | PT-004 | 50-54 | M | NEURO ICU | IPD | Dec 2025 | Urine (MSU) | Candida guilliermondii | ≥10^5 | Fungi | Automated ID & Sensitivity | 6557 |
| 5 | PT-005 | 60-64 | F | RESPIRATORY ICU | IPD | Dec 2025 | Urine (MSU) | Escherichia coli | ≥10^5 | Gram-negative | Automated ID & Sensitivity | 6344 |
| 6 | PT-006 | 60-64 | M | EMERGENCY ICU | IPD | Dec 2025 | Urine (MSU) | Escherichia coli | ≥10^5 | Gram-negative | Automated ID & Sensitivity | 6742 |
| 7 | PT-007 | 70-74 | F | MICU-I | IPD | Aug 2025 | Urine (MSU) | Escherichia coli | ≥10^5 | Gram-negative | Automated ID & Sensitivity | 3814 |
| 8 | PT-008 | 40-44 | M | RESPIRATORY ICU | IPD | Dec 2025 | Urine (MSU) | Candida lusitaniae | ≥10^5 | Fungi | Automated ID & Sensitivity | 6848 |
| 9 | PT-009 | 75-79 | F | DELUXE ROOMS | OPD | Dec 2025 | Urine (MSU) | Escherichia coli | ≥10^5 | Gram-negative | Automated ID & Sensitivity | 5198 |
| 10 | PT-010 | 50-54 | F | EMERGENCY ICU | OPD | Dec 2025 | Urine (MSU) | Escherichia coli | ≥10^5 | Gram-negative | Automated ID & Sensitivity | 3147 |
| 11 | PT-011 | 70-74 | M | OPD | OPD | Dec 2025 | Urine (MSU) | Escherichia coli | ≥10^5 | Gram-negative | Automated ID & Sensitivity | 3299 |
| 12 | PT-012 | 70-74 | M | OPD | OPD | Dec 2025 | Urine (MSU) | Escherichia coli | ≥10^5 | Gram-negative | Automated ID & Sensitivity | 3314 |
| 13 | PT-013 | 70-74 | M | CASUALTY WARD | IPD | Nov 2025 | Urine (MSU) | Escherichia coli | ≥10^5 | Gram-negative | Automated ID & Sensitivity | 5509 |
| 14 | PT-014 | 70-74 | F | EMERGENCY ICU | IPD | Nov 2025 | Urine (MSU) | Klebsiella pneumoniae | ≥10^5 | Gram-negative | Automated ID & Sensitivity | 5515 |
| 15 | PT-015 | 15-19 | F | SURGERY WARD IV | OPD | Nov 2025 | Urine (MSU) | Klebsiella pneumoniae | ≥10^5 | Gram-negative | Automated ID & Sensitivity | 6111 |
| 16 | PT-016 | 35-39 | F | OPD | OPD | Nov 2025 | Urine (MSU) | Enterococcus faecalis | ≥10^5 | Gram-positive | Automated ID & Sensitivity | 2749 |
| 17 | PT-017 | 45-49 | M | OPD | OPD | Nov 2025 | Urine (MSU) | Pseudomonas aeruginosa | ≥10^5 | Gram-negative | Automated ID & Sensitivity | 2790 |
| 18 | PT-018 | 85-89 | M | MICU-I | OPD | Nov 2025 | Urine (MSU) | Escherichia coli | ≥10^5 | Gram-negative | Automated ID & Sensitivity | 6209 |
| 19 | PT-019 | 85-89 | M | OPD | OPD | Nov 2025 | Urine (MSU) | Klebsiella pneumoniae | ≥10^5 | Gram-negative | Automated ID & Sensitivity | 2592 |
| 20 | PT-020 | 35-39 | M | GEN.MED ICU-2 | IPD | Nov 2025 | Urine (MSU) | Enterococcus faecium | ≥10^5 | Gram-positive | Automated ID & Sensitivity | 6178 |
| 21 | PT-021 | 35-39 | F | MEDICINE WARD-III | IPD | Nov 2025 | Urine (MSU) | Klebsiella pneumoniae | ≥10^5 | Gram-negative | Automated ID & Sensitivity | 6311 |
| 22 | PT-022 | 35-39 | F | NEURO ICU | IPD | Nov 2025 | Urine (MSU) | Klebsiella pneumoniae | ≥10^5 | Gram-negative | Automated ID & Sensitivity | 6735 |
| 23 | PT-023 | 35-39 | F | CASUALTY WARD | IPD | Nov 2025 | Urine (MSU) | Klebsiella pneumoniae | ≥10^5 | Gram-negative | Automated ID & Sensitivity | 6324 |
| 24 | PT-024 | 45-49 | F | EMERGENCY ICU | IPD | Sep 2025 | Urine (MSU) | Candida tropicalis | ≥10^5 | Fungi | Automated ID & Sensitivity | 6500 |
| 25 | PT-025 | 55-59 | F | RESPIRATORY ICU | IPD | Nov 2025 | Urine (MSU) | Candida albicans | ≥10^5 | Fungi | Automated ID & Sensitivity | 5765 |
| 26 | PT-026 | 65-69 | M | GEN.MED ICU-2 | IPD | Nov 2025 | Urine (MSU) | Enterobacter clocae | ≥10^5 | Gram-negative | Automated ID & Sensitivity | 5771 |
| 27 | PT-027 | 45-49 | F | MICU-I | IPD | Nov 2025 | Urine (MSU) | Enterococcus faecalis | ≥10^5 | Gram-positive | Automated ID & Sensitivity | 6039 |
| 28 | PT-028 | 55-59 | M | EMERGENCY HDU | IPD | Nov 2025 | Urine (MSU) | Escherichia coli | ≥10^5 | Gram-negative | Automated ID & Sensitivity | 6045 |
| 29 | PT-029 | 40-44 | F | EFEGENCY HDU | IPD | Oct 2025 | Urine (MSU) | Escherichia coli | ≥10^5 | Gram-negative | Automated ID & Sensitivity | 6870 |
| 30 | PT-030 | 55-59 | M | EMERGENCY ICU | IPD | Oct 2025 | Urine (MSU) | Klebsiella pneumoniae | ≥10^5 | Gram-negative | Automated ID & Sensitivity | 6903 |
| 31 | PT-031 | 70-74 | F | EMERGENCY ICU | IPD | Oct 2025 | Urine (MSU) | Klebsiella pneumoniae | ≥10^5 | Gram-negative | Automated ID & Sensitivity | 6918 |
| 32 | PT-032 | 50-54 | M | EMERGENCY ICU | IPD | Oct 2025 | Urine (MSU) | Klebsiella pneumoniae | ≥10^5 | Gram-negative | Automated ID & Sensitivity | 6957 |
| 33 | PT-033 | 75-79 | M | EMERGENCY ICU | IPD | Oct 2025 | Urine (MSU) | Escherichia coli | ≥10^5 | Gram-negative | Automated ID & Sensitivity | 6982 |
| 34 | PT-034 | 75-79 | M | EMERGENCY ICU | IPD | Jul 2025 | Urine (MSU) | Escherichia coli | ≥10^5 | Gram-negative | Automated ID & Sensitivity | 7035 |
| 35 | PT-035 | 95-99 | M | GEN.MED ICU-2 | IPD | Oct 2025 | Urine (MSU) | Enterococcus faecalis | ≥10^5 | Gram-positive | Automated ID & Sensitivity | 7047 |
| 36 | PT-036 | 60-64 | M | EMERGENCY ICU | IPD | Oct 2025 | Urine (MSU) | Enterobacter clocae | ≥10^5 | Gram-negative | Automated ID & Sensitivity | 7190 |
| 37 | PT-037 | 60-64 | F | EMERGENCY ICU | IPD | Oct 2025 | Urine (MSU) | Escherichia coli | ≥10^5 | Gram-negative | Automated ID & Sensitivity | 7197 |
| 38 | PT-038 | 75-79 | F | GEN.MED ICU-2 | IPD | Oct 2025 | Urine (MSU) | Klebsiella pneumoniae | ≥10^5 | Gram-negative | Automated ID & Sensitivity | 7253 |
| 39 | PT-039 | 65-69 | M | EMERGENCY HDU | IPD | Sep 2025 | Urine (MSU) | Escherichia coli | ≥10^5 | Gram-negative | Automated ID & Sensitivity | 7224 |
| 40 | PT-040 | 70-74 | F | EMERGENCY ICU | IPD | Sep 2025 | Urine (MSU) | Escherichia coli | ≥10^5 | Gram-negative | Automated ID & Sensitivity | 7291 |
| 41 | PT-041 | 45-49 | F | PICU-II | IPD | Sep 2025 | Urine (MSU) | Candida albicans | ≥10^5 | Fungi | Automated ID & Sensitivity | 7342 |
| 43 | PT-042 | 65-69 | F | OPD | OPD | Aug 2025 | Urine (MSU) | Klebsiella pneumoniae | ≥10^5 | Gram-negative | Automated ID & Sensitivity | 1929 |
| 47 | PT-043 | 25-29 | F | OPD | OPD | Jul 2025 | Urine (MSU) | Escherichia coli | ≥10^5 | Gram-negative | Automated ID & Sensitivity | 1777 |
| 44 | PT-044 | 55-59 | F | EMERGENCY ICU | IPD | Aug 2025 | Urine (MSU) | Pseudomonas aeruginosa | ≥10^5 | Gram-negative | Automated ID & Sensitivity | 4293 |
| 48 | PT-045 | 50-54 | M | GEN.MED ICU-2 | IPD | Jul 2025 | Urine (MSU) | Escherichia coli | ≥10^5 | Gram-negative | Automated ID & Sensitivity | 3768 |
| 42 | PT-046 | 20-24 | M | OPD | OPD | Jul 2025 | Urine (MSU) | Escherichia coli | ≥10^5 | Gram-negative | Automated ID & Sensitivity | 1632 |
| 46 | PT-047 | 70-74 | F | MICU-I | IPD | Jul 2025 | Urine (MSU) | Escherichia coli | ≥10^5 | Gram-negative | Automated ID & Sensitivity | 3708 |
| 45 | PT-048 | 55-59 | M | EMEGERNCY ICU | OPD | Jul 2025 | Urine (MSU) | Escherichia coli | ≥10^5 | Gram-negative | Automated ID & Sensitivity | 3635 |
| 49 | PT-049 | 25-29 | M | NEURO ICU | IPD | Aug 2025 | Urine (MSU) | Klebsiella pneumoniae | ≥10^5 | Gram-negative | Automated ID & Sensitivity | 4388 |
| 50 | PT-050 | 70-74 | F | EMEGERNCY ICU | IPD | Aug 2025 | Urine (MSU) | Candida albicans | ≥10^5 | Funi | Automated ID & Sensitivity | 4382 |

| S.No | Fosfomycin | Amoxicillin-Clav | Ampicillin | Ceftriaxone | Ciprofloxacin | Cotrimoxazole | Nitrofurantoin | Pip-Tazobactam | Amikacin | Cefepime | Colistin | Ertapenem | Gentamicin | Imipenem | Meropenem | Aztreonam | Piperacillin | Ceftazidime | Tetracycline |
|---|---|---|---|---|---|---|---|---|---|---|---|---|---|---|---|---|---|---|---|
| 1 | S | I | R | R | S | S | S | S | S | R | S | S | S | S | S | ND | ND | ND | ND |
| 2 | ND | ND | ND | ND | R | ND | ND | R | R | R | S | ND | ND | R | R | S | R | R | ND |
| 3 | S | R | R | R | R | R | R | R | I | R | S | R | R | R | R | ND | ND | ND | ND |
| 4 | ND | ND | ND | ND | ND | ND | ND | ND | ND | ND | ND | ND | ND | ND | ND | ND | ND | ND | ND |
| 5 | S | R | R | R | R | R | R | R | I | R | R | R | R | R | R | ND | ND | ND | ND |
| 6 | R | R | R | R | R | R | R | R | R | R | I | R | R | R | R | ND | ND | ND | ND |
| 7 | S | R | R | R | R | R | R | R | S | R | S | R | S | R | R | ND | ND | ND | ND |
| 8 | ND | ND | ND | ND | ND | ND | ND | ND | ND | ND | ND | ND | ND | ND | ND | ND | ND | ND | ND |
| 9 | S | R | R | R | R | R | R | S | S | R | RA | S | S | S | S | ND | ND | ND | ND |
| 10 | R | R | R | R | R | R | S | R | R | R | S | R | R | R | R | ND | ND | ND | ND |
| 11 | S | R | R | R | R | S | S | R | R | R | S | R | R | R | R | ND | ND | ND | ND |
| 12 | S | I | R | R | R | S | ND | R | S | R | RA | S | S | S | R | ND | ND | ND | ND |
| 13 | S | R | R | R | R | S | S | R | R | R | S | ND | R | S | R | ND | ND | ND | ND |
| 14 | R | R | R | R | R | R | R | R | R | R | S | R | R | R | R | ND | ND | ND | ND |
| 15 | S | R | R | R | R | R | R | R | R | R | S | R | R | R | R | ND | ND | ND | ND |
| 16 | R | ND | S | ND | R | ND | S | ND | ND | ND | ND | ND | ND | ND | ND | ND | ND | ND | R |
| 17 | ND | ND | ND | ND | R | R | ND | R | I | I | S | ND | ND | R | R | ND | ND | I | ND |
| 18 | ND | R | R | ND | R | R | ND | R | R | R | S | ND | R | R | R | R | ND | R | ND |
| 19 | ND | R | R | R | R | R | S | R | R | R | I | R | R | R | R | ND | ND | ND | ND |
| 20 | R | ND | ND | ND | R | ND | R | ND | ND | ND | ND | ND | ND | ND | ND | ND | R | ND | R |
| 21 | S | R | R | R | S | R | R | R | S | R | S | I | S | R | R | ND | ND | ND | ND |
| 22 | ND | R | R | R | R | R | ND | R | R | R | S | R | R | R | R | ND | ND | ND | ND |
| 23 | ND | R | ND | R | R | R | ND | R | R | R | S | R | R | R | R | R | ND | ND | ND |
| 24 | ND | ND | ND | ND | ND | ND | ND | ND | ND | ND | ND | ND | ND | ND | ND | ND | ND | ND | ND |
| 25 | ND | ND | ND | ND | ND | ND | ND | ND | ND | ND | ND | ND | ND | ND | ND | ND | ND | ND | ND |
| 26 | R | ND | ND | ND | R | R | ND | R | R | R | IR | ND | R | R | R | ND | ND | ND | ND |
| 27 | R | ND | S | ND | R | ND | S | ND | ND | ND | ND | ND | ND | ND | ND | ND | ND | ND | R |
| 28 | S | R | R | R | R | S | R | S | S | R | RA | S | R | S | S | ND | ND | ND | ND |
| 29 | S | R | R | R | R | R | R | R | R | R | S | R | R | R | R | ND | ND | ND | ND |
| 30 | ND | R | R | R | R | R | ND | R | R | R | S | R | R | R | R | ND | ND | ND | ND |
| 31 | R | R | R | R | R | R | R | R | R | R | S | R | R | R | R | ND | ND | ND | ND |
| 32 | ND | R | R | R | R | R | ND | R | R | R | S | R | R | R | R | ND | ND | ND | ND |
| 33 | S | I | R | R | R | R | S | R | I | R | S | S | S | R | R | ND | ND | ND | ND |
| 34 | S | R | R | R | R | R | R | R | R | R | S | R | R | R | R | ND | ND | ND | ND |
| 35 | R | ND | S | ND | R | ND | S | ND | ND | ND | ND | ND | ND | ND | ND | ND | ND | ND | R |
| 36 | R | R | R | ND | R | R | R | R | R | R | S | ND | R | R | R | R | ND | R | ND |
| 37 | S | R | R | ND | R | R | R | R | R | R | S | ND | R | R | R | R | ND | R | ND |
| 38 | ND | R | R | R | R | R | ND | R | R | R | S | R | R | R | R | ND | ND | ND | ND |
| 39 | S | R | R | R | R | R | S | R | R | R | S | R | R | R | R | ND | ND | ND | ND |
| 40 | R | R | R | ND | R | S | S | R | R | R | RA | ND | R | R | R | R | ND | R | ND |
| 41 | ND | ND | ND | ND | ND | ND | ND | ND | ND | ND | ND | ND | ND | ND | ND | ND | ND | ND | ND |
| 43 | ND | R | ND | R | R | R | ND | R | R | I | R | S | R | S | S | ND | ND | ND | ND |
| 47 | S | R | R | ND | R | ND | S | S | S | S | R | ND | S | S | S | S | ND | S | ND |
| 44 | ND | ND | ND | ND | R | ND | ND | I | R | I | R | ND | ND | S | I | ND | ND | R | ND |
| 48 | S | R | R | R | R | R | R | R | R | R | S | R | R | R | R | R | R | ND | ND |
| 42 | S | I | R | R | R | R | S | S | S | R | S | S | S | S | S | S | ND | ND | ND |
| 46 | S | ND | R | ND | R | R | R | R | S | R | S | ND | S | R | R | R | ND | R | ND |
| 45 | S | I | R | R | R | S | S | S | S | R | S | S | S | S | S | ND | ND | ND | ND |
| 49 | ND | R | ND | R | R | S | ND | R | R | R | S | R | R | I | R | ND | ND | ND | ND |
| 50 | ND | ND | ND | ND | ND | ND | ND | ND | ND | ND | ND | ND | ND | ND | ND | ND | ND | ND | ND |

| S.No | Amphotericin B | Caspofungin | Fluconazole | Micafungin | Voriconazole | Vancomycin | Linezolid | Teicoplanin | Penicillin G | High-Level Gentamicin | ESBL Screening | Carbapenemase Screening | MDR (Auto) | ESBL+ Flag | CRE Flag |
|---|---|---|---|---|---|---|---|---|---|---|---|---|---|---|---|
| 1 | ND | ND | ND | ND | ND | ND | ND | ND | ND | ND | Positive | Negative | YES | ESBL+ | — |
| 2 | ND | ND | ND | ND | ND | ND | ND | ND | ND | ND | ND | Negative | YES | — | — |
| 3 | ND | ND | ND | ND | ND | ND | ND | ND | ND | ND | Positive | Positive | YES | ESBL+ | CRE |
| 4 | S | S | S | S | S | ND | ND | ND | ND | ND | ND | ND | NO | — | — |
| 5 | ND | ND | ND | ND | ND | ND | ND | ND | ND | ND | Positive | Positive | YES | ESBL+ | CRE |
| 6 | ND | ND | ND | ND | ND | ND | ND | ND | ND | ND | Positive | Positive | YES | ESBL+ | CRE |
| 7 | ND | ND | ND | ND | ND | ND | ND | ND | ND | ND | Positive | Positive | YES | ESBL+ | CRE |
| 8 | S | ND | S | ND | S | ND | ND | ND | ND | ND | ND | ND | NO | — | — |
| 9 | ND | ND | ND | ND | ND | ND | ND | ND | ND | ND | Positive | Negative | YES | ESBL+ | — |
| 10 | ND | ND | ND | ND | ND | ND | ND | ND | ND | ND | Positive | Positive | YES | ESBL+ | CRE |
| 11 | ND | ND | ND | ND | ND | ND | ND | ND | ND | ND | Positive | Positive | YES | ESBL+ | CRE |
| 12 | ND | ND | ND | ND | ND | ND | ND | ND | ND | ND | Positive | Positive | YES | ESBL+ | CRE |
| 13 | ND | ND | ND | ND | ND | ND | ND | ND | ND | ND | Positive | Positive | YES | ESBL+ | CRE |
| 14 | ND | ND | ND | ND | ND | ND | ND | ND | ND | ND | Positive | Positive | YES | ESBL+ | CRE |
| 15 | ND | ND | ND | ND | ND | ND | ND | ND | ND | ND | Positive | Positive | YES | ESBL+ | CRE |
| 16 | ND | ND | ND | ND | ND | ND | S | S | R | ND | ND | ND | YES | — | — |
| 17 | ND | ND | ND | ND | ND | ND | ND | ND | ND | ND | ND | Positive | YES | — | CRE |
| 18 | ND | ND | ND | ND | ND | ND | ND | ND | ND | ND | Positive | Positive | YES | ESBL+ | CRE |
| 19 | ND | ND | ND | ND | ND | ND | ND | ND | ND | ND | Positive | Positive | YES | ESBL+ | CRE |
| 20 | ND | ND | ND | ND | ND | R | S | R | R | R | ND | ND | YES | — | — |
| 21 | ND | ND | ND | ND | ND | ND | ND | ND | ND | ND | Positive | Positive | YES | ESBL+ | CRE |
| 22 | ND | ND | ND | ND | ND | ND | ND | ND | ND | ND | Positive | Positive | YES | ESBL+ | CRE |
| 23 | ND | ND | ND | ND | ND | ND | ND | ND | ND | ND | Positive | Positive | YES | ESBL+ | CRE |
| 24 | S | S | S | S | S | ND | ND | ND | ND | ND | ND | ND | NO | — | — |
| 25 | S | S | S | S | S | ND | ND | ND | ND | ND | ND | ND | NO | — | — |
| 26 | ND | ND | ND | ND | ND | ND | ND | ND | ND | ND | Positive | Positive | YES | ESBL+ | CRE |
| 27 | ND | ND | ND | ND | ND | S | S | S | R | R | ND | ND | YES | — | — |
| 28 | ND | ND | ND | ND | ND | ND | ND | ND | ND | ND | Positive | Positive | YES | ESBL+ | CRE |
| 29 | ND | ND | ND | ND | ND | ND | ND | ND | ND | ND | Positive | Positive | YES | ESBL+ | CRE |
| 30 | ND | ND | ND | ND | ND | ND | ND | ND | ND | ND | Positive | Positive | YES | ESBL+ | CRE |
| 31 | ND | ND | ND | ND | ND | ND | ND | ND | ND | ND | Positive | Positive | YES | ESBL+ | CRE |
| 32 | ND | ND | ND | ND | ND | ND | ND | ND | ND | ND | Positive | Positive | YES | ESBL+ | CRE |
| 33 | ND | ND | ND | ND | ND | ND | ND | ND | ND | ND | Positive | Positive | YES | ESBL+ | CRE |
| 34 | ND | ND | ND | ND | ND | ND | ND | ND | ND | ND | Positive | Positive | YES | ESBL+ | CRE |
| 35 | ND | ND | ND | ND | ND | S | S | S | S | R | ND | ND | YES | — | — |
| 36 | ND | ND | ND | ND | ND | ND | ND | ND | ND | ND | Positive | Positive | YES | ESBL+ | CRE |
| 37 | ND | ND | ND | ND | ND | ND | ND | ND | ND | ND | Positive | Positive | YES | ESBL+ | CRE |
| 38 | ND | ND | ND | ND | ND | ND | ND | ND | ND | ND | Positive | Positive | YES | ESBL+ | CRE |
| 39 | ND | ND | ND | ND | ND | ND | ND | ND | ND | ND | Positive | Positive | YES | ESBL+ | CRE |
| 40 | ND | ND | ND | ND | ND | ND | ND | ND | ND | ND | Positive | Positive | YES | ESBL+ | CRE |
| 41 | S | S | S | S | S | ND | ND | ND | ND | ND | ND | ND | NO | — | — |
| 43 | ND | ND | ND | ND | ND | ND | ND | ND | ND | ND | Positive | Negative | YES | ESBL+ | — |
| 47 | ND | ND | ND | ND | ND | ND | ND | ND | ND | ND | Positive | Positive | YES | ESBL+ | CRE |
| 44 | ND | ND | ND | ND | ND | ND | ND | ND | ND | ND | ND | Negative | YES | — | — |
| 48 | ND | ND | ND | ND | ND | ND | ND | ND | ND | ND | Positive | Positive | YES | ESBL+ | CRE |
| 42 | ND | ND | ND | ND | ND | ND | ND | ND | ND | ND | Positive | Negative | YES | ESBL+ | — |
| 46 | ND | ND | ND | ND | ND | ND | ND | ND | ND | ND | Positive | Positive | YES | ESBL+ | CRE |
| 45 | ND | ND | ND | ND | ND | ND | ND | ND | ND | ND | Positive | Positive | YES | ESBL+ | CRE |
| 49 | ND | ND | ND | ND | ND | ND | ND | ND | ND | ND | Positive | Positive | YES | ESBL+ | CRE |
| 50 | S | S | S | S | S | ND | ND | ND | ND | ND | ND | ND | NO | — | — |

### ANTIBIOGRAM — Antimicrobial Susceptibility Profiles of Major Uropathogens

*Chhatrapati Shivaji Subharti Hospital, Meerut · Six-Month Retrospective Study*

| Susceptible (S%) | Intermediate (I%) | Resistant (R%) | Not Tested — NA |
|---|---|---|---|

#### Gram Negative Rods (GNR)

| Organism | | Colistin | Fosfomycin | Nitrofurantoin | Cotrimoxazole | Ciprofloxacin | Gentamicin | Amikacin | Meropenem | Imipenem | Ertapenem | Cefepime | Ceftriaxone | Ceftazidime | Pip-Tazo | Amox-Clav | Ampicillin |
|---|---|---|---|---|---|---|---|---|---|---|---|---|---|---|---|---|---|
| **E. coli** *n = 23* | S% | 84.2 | 86.4 | 47.6 | 31.8 | 4.3 | 39.1 | 39.1 | 26.1 | 34.8 | 41.2 | 4.3 | 0 | NA | 26.1 | 0 | 0 |
| | I% | 5.3 | 0 | 0 | 0 | 0 | 0 | 13.0 | 0 | 0 | 0 | 0 | 0 | NA | 0 | 22.7 | 0 |
| | R% | 10.5 | 13.6 | 52.4 | 68.2 | 95.7 | 60.9 | 47.8 | 73.9 | 65.2 | 58.8 | 95.7 | 100 | NA | 73.9 | 77.3 | 100 |
| **K. pneumoniae** *n = 12* | S% | 83.3 | 50 | 20 | 8.3 | 8.3 | 8.3 | 8.3 | 8.3 | 8.3 | 8.3 | 0 | 0 | NA | 0 | 0 | NA |
| | I% | 8.3 | 0 | 0 | 0 | 0 | 0 | 0 | 0 | 8.3 | 8.3 | 8.3 | 0 | NA | 0 | 0 | NA |
| | R% | 8.3 | 50 | 80 | 91.7 | 91.7 | 91.7 | 91.7 | 91.7 | 83.3 | 83.3 | 91.7 | 100 | NA | 100 | 100 | NA |
| **P. aeruginosa** *n = 3* | S% | 66.7 | NA | NA | NA | 0 | NA | 0 | 0 | 33.3 | NA | 0 | NA | 0 | 0 | NA | NA |
| | I% | 0 | NA | NA | NA | 0 | NA | 33.3 | 33.3 | 0 | NA | 66.7 | NA | 33.3 | 33 | NA | NA |
| | R% | 33.3 | NA | NA | NA | 100 | NA | 66.7 | 66.7 | 66.7 | NA | 33.3 | NA | 66.7 | 66.7 | NA | NA |

#### Gram Positive Cocci / Enterococcus (GPC)

| Organism | | HL Gentamicin | Linezolid | Teicoplanin | Vancomycin | Tetracycline | Nitrofurantoin | Fosfomycin | Ciprofloxacin | Penicillin G | Ampicillin |
|---|---|---|---|---|---|---|---|---|---|---|---|
| **E. faecalis** *n = 3* | S% | 0 | 100 | 100 | 100 | 0 | 100 | 0 | 0 | 33.3 | 0 |
| | I% | 0 | 0 | 0 | 0 | 0 | 0 | 0 | 0 | 0 | 0 |
| | R% | 100 | 0 | 0 | 0 | 100 | 0 | 100 | 100 | 66.7 | 100 |

**Note:** *S = Susceptible; I = Intermediate; R = Resistant; NA = Not Tested. Values are percentages of isolates. Testing per CLSI/EUCAST breakpoints.*
